# Developing a culturally tailored brain healthy diet intervention for older midwestern African Americans: A Qualitative and Quantitative Study

**DOI:** 10.1101/2022.06.02.22275914

**Authors:** Ashley R. Shaw, Eric D. Vidoni, Debra K. Sullivan, Jannette Berkley-Patton, Jeffrey Burns

## Abstract

**Background:** Alzheimer’s disease and related dementia (ADRD) in the United States (U.S.) disproportionately affects African Americans (AA). Previous research has indicated that healthy eating habits help slow cognitive decline among adults 65 years and older. However, dietary intervention studies that demonstrated preliminary ability to protect against cognitive decline demonstrated low adherence and acceptability among African Americans.

**Objective:** The objective of this study is to identify and understand knowledge and attitudes that influence dietary practices among older African Americans using a community-engaged approach.

**Design:** A non-interventional mixed methods study designed to inform the development of an adapted brain-healthy soul food diet intervention. A purposive sampling approach was used to conduct 5 semi-structured focus group discussions and an online quantitative survey. The Health Belief Model guided the focus group using the following constructs: susceptibility, perceived severity, perceived benefits, perceived barriers, self-efficacy, and cue to action.

**Participants/setting:** Inclusion criteria included self-identifying as African American or Black, aged 55 years and older, English proficient, and cognitively normal with an AD8 < 2. Participant were asked to participate in a single 60-minute virtual focus group discussion. In total, 39 individuals (25.6% men, 74.4% women) took part in one of the seven virtual focus group discussion (5-7 per focus group).

**Main outcome measure:** Knowledge and attitudes that influence dietary practices among older African Americans.

**Statistical analysis performed:** Focus groups were analyzed using a 6-step thematic analysis process, and quantitative survey data was analyzed using descriptive statistics.

**Results:** Five themes emerged: Knowledge of dementia; practices shaping food choices and consumption; barriers impacting healthy dietary consumption; experiential instrumental support; and elements of a culturally tailored brain healthy dietary intervention.

**Conclusion:** Older Midwesterner’s perceived an adapted MIND dietary model as most feasible with the incorporation of salient cultural characteristics and strategies within both the design and delivery phases.

**Research Snapshot:** *Research Question:* What are the perceptions related to healthy diets and the facilitators and barriers that influence dietary practices among older Midwestern African Americans?

*Key Finding:* Five themes emerged from the focus group discussions: Knowledge of dementia; Practices shaping food choices and consumption; Barriers impacting healthy dietary consumption; External instrumental support; and Elements of culturally tailored brain healthy dietary intervention. Facilitators identified to support following a healthy diet included cooking education, food preparation demonstrations, and accessibility guidance. Barriers identified that impact healthy dietary consumption included access, cost, taste, and food spoilage.

---

Alzheimer’s disease and related dementias (ADRD) in the United States (U.S.) disproportionately affect African Americans (AA). AAs are two to three times more likely to develop Alzheimer’s Disease (AD) compared to non-Hispanic Whites, with AD being the 4^th^ leading cause of death among AA.^1^

Previous research has indicated that healthy eating habits and requisite diets such as the Mediterranean diet, Dietary Approaches to Stop Hypertension (DASH), and Mediterranean-DASH Intervention for Neurogenerative Delay (MIND) help slow cognitive decline among adults 65 years and older.^2-4^ Large studies such as 1.5 year study including non-demented adults found that higher adherence to the Mediterranean diet was associated with up to a 40% reduced risk for AD.^3^ Another study found that adherence to the MIND diet was associated with slower cognitive decline and individual cognitive domains increased by 30-78%.^2^ However, much of the research regarding dietary prevention of AD is based almost exclusively on studies involving non-Hispanic Whites resulting in limited generalizability of findings for older adults who belong to underrepresented and understudied racial/ethnic groups.^2^

Dietary intervention studies that demonstrate preliminary or cursory ability to protect against cognitive decline, including among AAs, have demonstrated low adherence and acceptability due to perceived lack of social support, social contexts (i.e. cost of healthy foods, reduced access to healthy and diverse food options due to low socioeconomic status), strong cultural influence on food preferences and preparation, and perceived less appealing taste of low fat foods.^5-7^ Previous research has indicated that modifying soul food or traditional AA recipes to better meet nutritional guidelines would be more effective than suggesting that such foods be eliminated from a healthy diet altogether.^8^ For example, culturally adapted diets have resulted in AA participants lowering BMI, A1C, reducing body weight and waist circumference.^9,10^ It is important to note that traditional soul foods include healthy dietary components,^11^ such as fruits and vegetables (e.g. collard greens, sweet potatoes, okra, and blueberries), which are linked to improved cardiovascular risk markers and slower cognitive decline.^12,13^ Moreover, such dietary interventions developed for AA have been guided by community participation.^14^ These elements are consistent with our emerging understanding of what makes up a brain healthy diet.

Use of qualitative research allows for communities to reflect upon their experiences and beliefs as they relate to food and eating patterns in their own words. Qualitative research has been shown to be useful in advancing knowledge and understanding regarding aspects of food and eating practices.^15^ Focusing on attitudes, beliefs, practices, and the cultural meanings of health dietary patterns from the perspective of the community can provide context to future culturally appropriate dietary intervention strategies and inform policy approaches.^16^

Community engaged research (CER) is an approach that consciously involves community members impacted by a phenomenon by incorporating their insights throughout the research process which can support the development of interventions and influence policies.^17,18^ CER has been widely recognized in research as imperative to reduce health disparities.^19-21^ Use of CER within qualitative research can enhance translational results. Previous research has found that studies incorporating a CER approach has a positive impact on health behaviors including diet, exercise, and health outcomes (i.e. quality of life).^22^ Use of CER was well suited for this study’s development of a brain healthy diet intervention because it can address the intersection between current dietary practices with social determinates of health in a way that fosters a deeper understanding of culturally relevant concerns and can target multi-faceted barriers related to health.^23,24^

## Objective

The objective of this study is to identify and understand knowledge and attitudes that influence dietary practices among older African Americans using a community-engaged approach.

## METHODS

Approval to conduct the study was obtained by the University of Kansas Medical Center Institutional Review Board (IRB#00146894). This was a non-interventional mixed method study designed to inform the development of an adapted brain-healthy soul food diet intervention. Specifically, this study aimed to understand older AA specific food preferences, preparation preferences, and community cooking class topics specific to support the development and delivery of the future adapted brain healthy diet intervention. Additionally, this study aimed to understand barriers and facilitator of dietary behaviors among older AA to inform the development of the adapted bran-healthy soul food diet intervention.

### Participants and Recruitment

We used purposive sampling to recruit participants through existing registries, distribution of flyers, and by emails from the principal investigator (PI). Individuals interested in participating in the study contacted research staff by phone and completed a brief telephone screening to verify eligibility. Study eligibility requirements including self-identifying as AA or Black, aged 55 years and older, English proficient, and cognitively normal with an AD8 Dementia Screening of ≤ 2.

### Focus Group Questionnaire Development

The Health Belief Model (HBM) was used to guide the development of the semi-structured focus group questionnaire. Specifically, the focus group questions were guided by the following constructs of the HBM perceived susceptibility, perceived severity, perceived benefits, perceived barriers, self-efficacy, and cue to action^25^. The HBM was chosen because it has been widely used in diverse cultural context areas and within dietary-related intervention development^26-28^. The research team developed a pool of potential questions for the brain health and dietary discussion guide. From the pool, a total of 17 questions were selected in addition to follow-up probes.

### Procedure

#### Online Surveys

All participants provided written consent before participating in the study. Prior to focus group discussions 39 participants completed a 30 minute online demographic survey (15-item), eating habits survey (15-item), and needs assessment survey (26-item). The surveys included questions about demographics, health care, eating patterns, cultural food needs, and community nutrition education needs. The survey was administered through a REDCap online link that was emailed individually to each participant 7 days prior to their scheduled focus group discussion.

#### Focus Groups

Five focus group discussions, each consisting of 5-7 people (39 total), were conducted between May 2021 – July 2021 and each focus group took place virtually using the Zoom platform. Each focus group lasted 60 minutes. Focus groups followed a semi-structured guide in which open-ended, clear, and concise questions were used to stimulate discussion. Additionally, probe questions were used to obtain more detailed information from focus group participants. All focus group discussions were audio recorded and transcribed verbatim. The focus groups were led by a trained AA facilitator. She was selected to facilitate due to her education, prior experience moderating focus group discussions in the AA community, and cultural relation and understanding of the participants. Participants received a $50 gift card in appreciation for their time.

#### Member Check Focus Group

Upon completion of the data analysis (see below), we conducted member checking, or participant validation to ensure participants perspectives were appropriately represented and not curtailed by the researchers knowledge and personal agenda.^29^ The member check focus group was comprised of a subset of participants (n=5), was led by the AA facilitator, and took place 3 months after the final focus group discussion via Zoom platform. The member check focus group in this study was conducted to ensure that emerging themes were congruent with the community’s interpretation, resulting in a final thematic consensus. The single member check focus group lasted 30 minutes and gave participants an opportunity to check for accuracy of the data, which is consistent with best practices for exploring credibility and enhancing rigor of qualitative data^29,30^

### Data Analysis

#### Quantitative data

Demographics of participants and online surveys were analyzed using descriptive statistics.

#### Qualitative data

Transcribed data was placed into Dedoose, a qualitative analysis software program,^31^ which supported coding, filing, and organizing themes and subthemes. Data was analyzed using a 6-step thematic analysis process including familiarization, coding, generating themes, reviewing themes, defining, and naming themes, and write up.^32^

## RESULTS

A total of 39 people participated in the study. Most participants were female (74.4%); 65 years and older (60.3%), with an education level equivalent or higher to a bachelor’s degree (61.5%), retired (69.2%), who identified as a Christian (94.9%). Additionally, most participants reported having cardiovascular related health conditions such as hypertension (66.7%) or high cholesterol (51.3%). Additional demographic details about the sample are shown in Table 1.

### Online Survey Results

#### Eating habits

Most participants indicated that they currently either do not follow a dietary eating practice (48.7%) or follow a low sodium eating diet (25.6%). For food preparation most participants reported regularly baking (82.1%), grilling (46.2%), or air frying (43.6%) their food. A full description of reported eating habits can be found in Table 2.

#### Health conditions knowledge interest

Many participants indicated that they were “very interested” or “interested” in receiving more health education related to AD (89.7%) and blood pressure (76.9%) (Table 3).

#### Barriers to eating healthy

Regarding perceived barriers to eating healthy many participants indicated that taste (30.8%) a being one of the primary reasons followed by cost (20.5%). A full description of perceived barriers to eating healthy can be found in Table 4.

#### Nutrition education and delivery preferences

For a dietary intervention participants reported that they were either very interested or interested in receiving education related to water intake (94.9%), meal planning and prepping (87.2%), nutritional content (87.1%), and eating healthy on a budget (84.6%). Most people indicated that they preferred to support from a nutritionist or health coach virtually (94.9%) or by phone (94.9%).

Additionally, most people reported that they would prefer to receive a dietary intervention virtually (94.9%) or at a church (94.9%). See Table 5 for nutrition education and delivery preferences.

### Focus Group Discussion Themes

A total of 5 themes emerged from the focus group discussions. The results are summarized according to the major themes and subthemes expressed across all 7 focus group discussions. See table 6 for a summary of themes, subthemes, and sample quotes.

### Theme 1. Knowledge of dementia

All participant reported various ways of first learning about dementia. For theme 1 we identified 3 subthemes: family history, perceived cause of dementia, and role of diet in dementia.

#### Family history

Many participants expressed that they first learned about dementia from having a family history of dementia (i.e. grandparents, parents, in-laws, older siblings). *“I first learned about it with my father. My father was diagnosed with it and I just say if you’ve ever been diagnosed with dementia, might as well say you have Alzheimer’s. So that’s how I first learned about it that was about 16 or 17 years ago*.*”* Some participants also mentioned that they first learned about dementia through media outlets (i.e. articles, AARP) and through church in which older congregant members at their church had dementia.

#### Perceived cause of dementia

Participants across all focus group groups perceived dementia to be prevalent among older people in general not necessarily more prevalent among Black Americans compared to other racial/ethnic groups. However, several participants believed that dementia is serious in the Black community due to the lack of awareness the community has about what dementia is and the perception that it is a normal part of aging (i.e. senility). Participants voiced that they believed contributing factors to lack of awareness stemmed from lack of education and lack of access to available resources. *“I’m feeling like it is a serious problem for Black people…. we don’t have the access or the resources or sometimes the will to go and find out what is going on with us or with a relative. On the other hand, I would hesitate to say definitively that there’s this more serious for Black people ’cause I have not seen statistics on that*.*”*

#### Role of diet in dementia

Regarding the role of diet in dementia most participants believed that following a healthy diet was important for keeping the brain healthy. *“I think that a healthy diet is important for every part of your body, especially the brain, because if we give the brain the correct nutrients, I just believe that we will, you know, function better and function longer if we eat the right things*.*”* Specifically, participants perceived that regularly following a healthy diet, especially early in life can provided essential nutrients to keep the brain functioning healthy as we age. However, some participants voiced that they believed that healthy eating is not commonly practiced until one gets older, when people start thinking more about longevity of life.

### Theme 2. Practices shaping food choices and consumption

#### Household decision making

Many participants indicated that they lived alone therefore household decision making about food was made individually. *“I live with myself, so me*.*”* Among those who indicated that they were married many indicated that the wife did a majority of the decision-making regarding food. Lastly, the key factor that played a role within the decision making for food was cost.

#### Food purchasing practices

Participants voiced that food purchasing practices were largely influenced by cost and that they shopped based of deals from grocery-ads and deals in their supermarkets. Some participants indicated that due to the COVID-19 pandemic their food purchasing practices have shifted to primarily grocery shopping online, with the exception of going in-person to a grocery store to purchase produce. *“Since the pandemic, I have had like uh most of my shopping online and I go and pick it up, and I have a few smaller stores that I go to purchase things that um things like vegetables, things that I usually go in person to pick those out, not at a smaller store*.*”* Also, participants indicated that food purchasing practices including shopping for a variety of food within a week due to the preference of not wanting to eat the same foods/meals several times throughout the week.

#### Food preparation practices

Most participants voiced that they liked to use a variety of food preparation practices including baking, air frying, and grilling. It is important to note that many participants stated that frying foods was rarely practiced within their households. *“I sauté. I’m using the oven, as the weather continues to change, and now that I can make a good fire, I might use the grill more. But frying is just not something I do not do at all*.*”*

### Theme 3. Barriers impacting healthy dietary consumption

All participants across focus groups reported various barriers that impacted their ability to practice dietary consumption. For theme 3, we identified 4 subthemes: Access, Cost, Taste, and Food Spoilage.

#### Access

Many participants voiced access to healthy foods as being a significant barrier to eating healthy in the Black community. Specifically, many participants indicated that there are food deserts in many of the predominantly Black neighborhoods coupled with poor public transportation which makes it difficult to access healthy foods. *“In many of the communities they don’t have access to fresh fruits and vegetables…Places like when I’m walking is a food desert, you know they don’t have uh fresh produce in the stores in the neighborhoods, so if they don’t have it there, they can’t get it*.*”* Additionally, many participants stated that stores that are available in these neighborhoods do not offer fresh produce results in community members purchasing foods that are readily available, which often aren’t healthy options.

#### Cost

A majority of the participants across all focus groups voiced that cost of healthy food was a significant barrier to eating healthy in the Black community (i.e. fruits and vegetables) and believed that people would eat healthier if they could afford it. *“Cost. I think that, I think that if you eat healthier, it’s going to cost you more, instead of being an investment in your life, so some of the plant based foods are more expensive, and some of the plant based cheese are more expensive*.*”* Additionally, some participants believed cost was more of a barrier for people living in households with more than one generation. Overall cost of foods inhibits the Black community’s from being capable of fully investing in their health due to lack of affordability.

#### Taste

Some participants perceived taste as a barrier to consuming healthy food noting that healthy food often taste bland. Also, it was perceived that people in the Black community are more accustomed to foods that have higher sodium for seasoning which makes it less appetizing when consuming healthy foods of lower sodium. *“Sometimes people are used to the salt, and they don’t want to eat bland food*.*”*

#### Food Spoilage

Participants indicated that they perceived healthy foods, particularly produce to spoil quicky which hindered their ability to follow a healthy diet. *“I like vegetables, so it’s not a problem. The only problem is just keeping the vegetables on hand, you know without them spoiling and I have to throw them out*.*”* Specifically, people perceived fresh produce to have a short shelf life meaning that they would need to make multiple trips to the grocery store throughout the week, which can be burdensome.

### Theme 4. External instrumental support

The most important support mechanisms that were voiced across all focus groups to help people follow a healthy diet include education and accessibility guidance.

#### Cooking Education

Across all focus group participants highlighted the importance of providing cooking education to support buy-in from the community. *“I just think, more education. If you go out in the community and maybe even do demonstrations where you can show people that, how the food tastes, how easy it is to prepare*.*”* Many participants believed that education centers around cooking demonstrations as a means to explore new foods. Cooking demonstrations coupled with sample tasting allows for the community to get a better sense of healthy foods that they may like/don’t like so that they can incorporate more healthy foods in their diet moving forward.

#### Accessibility Guidance

Many participants indicated that although providing education (i.e. recipes, cooking demonstrations) is helpful in providing support to a more healthy dietary lifestyle in the Black community it is imperative to show where and how people can obtain healthy foods (i.e. highlighting specific grocery stores, farmers markets). *“It also has to be available, we can educate, we can show, we can even provide taste, but we have to make it available and show them where they can obtain or acquire these healthy foods, these recipes, these meals*.*”*

### Theme 5. Elements of culturally tailored brain healthy dietary intervention

All focus groups discussed ideas about the design of a brain healthy dietary intervention with the goal of improving healthy eating in the Black community to reduce the risk of AD. As a result, participants emphasized the importance of researchers incorporating a comprehensive recruitment and delivery method, use of an adapted MIND diet model, and emphasizing benefits to the community to support retention efforts.

#### Recruitment

Several people stated the recruitment into a brain healthy diet intervention needs to be comprehensive, meaning that it should include diverse outlets to reach a wide range of diverse older adults (across the city) within the Black community. Specifically, recruitment through media outlets such as the newspapers, radio, and tv ads would be best in reaching older Black adults. *“It needs to be put out comprehensively. Television, radio, magazines, newspapers, social media, churches, groups go into nursing homes and things of that nature that’s you know, a comprehensive approach because you can’t just utilize one level of approach. Because that’s only going to affect only a segment of that community*.*”* Many participants across focus groups also voiced the importance of establishing partnerships local churches and community centers who could support recruitment efforts by distributing recruitment materials via newsletter and email. Notably, social media outlets (i.e. Facebook and Twitter) were not favorably viewed by many of the participants as a way to recruit older adults due to irregular use of the platforms.

#### MIND Dietary Model

Of the 3 diets (MIND, Mediterranean, and DASH) discussed at each of the focus group sessions the MIND diet model was most favored and seen most feasible to follow due to the variety of food options and because it was viewed as aligning more with lifestyle practices within the black community. *“I like the variety; you know most of it, I’ve kind of been doing already. Not much of the olive oil, like I said I stick with kind of canola oil. So, it limits you on the cheese, I see that. I mean I use way more cheese than one serving a week. Everything else is OK*.*”* Moreover, the MIND diet was viewed as less restrictive compared to the Mediterranean diet. However, many participants voiced that some adjustments in the model are needed to increase acceptability within the Black community including increasing the recommended amount of butter and cheese as well as providing plant-based alternatives options in these categories. *“Less than, 1…if I’m fixing broccoli, if I’m fixing potatoes, they’re all gonna have some type of butter in it. If I’m fixing corn, it has butter in it. So, I don’t know if this is do-able for me*.*”* Participants also voiced the importance of including information about spices within the model to address the taste barrier many people in the Black community have. *“There is a vaster range of herbs and spices out there and we have to learn to use those”* Several participants discussed how they felt berries were not necessarily aligned with Black culture so alternative fruits that support brain health that are affordable should be highlighted such as cantaloupe which is high in antioxidants.^33^ *“I don*’*t like certain textures, so I don*’*t like blueberries or berries for that reason. But I could mix them into something for the flavor*.*” “I think I like it even more if I can eat cantaloupe…berries cost a little bit more than cantaloupe and apples and things like that*.*”* Additionally, more specific information about serving sizes in lay language need to be included so it is easier for people to follow. Many focus group participants believed it was imperative to include information about the importance of water intake in relation to the MIND diet. For the adapted MIND diet model participants voiced the importance of including foods for each of the 10 categories that the Black community would easily recognize (i.e. green leafy vegetables - collard greens, starchy vegetable – sweet potato, whole grains - oatmeal). Lastly, for each of the 10 food categories on the MIND diet many participants indicated that they felt that it was important to include a snippet of health information (i.e. sweet potatoes contain large amounts of fiber which improve the health of the gut and digestion) so people can understand additional benefits food groups have for the entire body, not just benefits for brain health.

#### Comprehensive Education and Delivery Method

Many focus group participants voiced the importance of a dietary intervention including a faith component (i.e. scriptures, health devotionals) coupled with ongoing education with resources (i.e. cooking classes with samples, simple recipes, preparation videos, pocket cards (MIND diet).

> *“I know being a person of faith here at, being with my church, there are scriptures that talks about taking back what the enemy has stole from you. So that includes your mind. We often go into material things, but your mind and your health is something that it was stolen because we did not eat correctly because of lack of education and resources. So, if they are available, you know for me, myself, I would use scripture going right along with the MIND diet. So, you prayed this scripture in the natural, what are you going to……. I mean for the spiritual piece, what are you going to do in the natural? So, in the natural pray this scripture, and in the physical eat the food along with it and walk along with it and let’s see what’s going to come. So, let’s just be you know, realistic with that as far as myself, that is the way that I’m going to approach the seniors here at our church about this MIND diet so we’re praying this, let’s put this into action*.*”*

Regarding delivery of education, participants across all focus groups indicated the importance of offering a hybrid model which would provide greater access to receiving education in the Black community. Additionally, for in-person education many participants believed it should be offered in community settings such as community centers and recreational centers.

#### Retention

To support retention efforts in the brain healthy diet intervention many participants voiced that it would be helpful to provide the community with various incentives (i.e. financial incentives such as grocery store gift cards to off-set the cost of healthy food). Since cognitive changes take time, many participants voiced the importance of highlighting short term benefits that will take place by following a healthy diet. Specifically, several participants emphasized the importance of designing an intervention that highlights the early benefits of following a healthy diet (i.e. weight loss, lower blood pressure, potential financial benefits from getting off medication), which would result in a higher retention rate in a brain healthy diet intervention. *“Incentives might work too, if people stay on this and they know they’ve lost some weight, maybe a small token or reward uh you know, or a trip or free meal, I don*’*t know, something. Incentives always help people*.*”*

## Discussion

The aim of this mixed methods study was to identify and understand knowledge and attitudes that influence dietary practices among older AAs using a community-engaged approach. More specifically, our study examined knowledge and attitudes that influence dietary practices to inform the development of an adapted brain-healthy soul food diet intervention. Overall, our findings indicated that older AAs perceive healthy dietary consumption to be imperative in terms of obtaining brain health. However, healthy dietary consumption is largely influenced by cultural attitudes, unhealthy patterns, social and environmental determinants which can inhibits one from achieving optimal health. Prior research suggests unhealthy dietary consumption among older adults is associated with several factors including income, lower education, social isolation, and inadequate detention, which emphasizes the role social determinants have in healthy behavior.^34^ In line with previous research^35^, our focus groups highlighted barriers to healthy dietary consumption which were largely influenced by disparities in access, availability, and cost of healthy foods.

This study adds to the limited research by providing context to the attitudes and beliefs shaping current dietary practices and provide information regarding culturally specific needs related to brain healthy dietary interventions for older AAs. Findings indicate that adapting the MIND diet to community settings through culturally appropriate food guidelines and practices can support buy-in by older Midwestern AAs. Previous research has indicated that AA associate specific foods, cooking techniques, and seasonings with their identity as an AA^6,11^, which aligns with findings from this study in which participants voiced the importance of focusing on foods aligned with the MIND diet that are well known within the AA community as well as sharing information regarding spices and herbs to regularly incorporate within an adapted diet. Adding to previous literature, highlighting traditional healthy soul foods and practices to an adapted MIND diet intervention is imperative in terms of having significant meaning related to passing on family traditions which in turn provides a sense of familiarity and respect to one’s culture.

Previous research has indicated the importance of including approaches that resonate with cultural and learning styles when developing and delivering interventions. Specifically, a holistic intervention approach that highlights the mind, body, spirit connection has been found to be essential to support reducing health disparities risk among AAs^36,37^. Our study adds to this literature in which findings from the study emphasized the importance of incorporating interconnectedness within the design of the adapted MIND diet educational curriculum.

### Strengths and Limitations

#### Strengths

Our study has several strengths including use of the mixed methods approach, resulting in a more thorough knowledge imperative to inform future intervention development. Also, use of focus groups provided a richer and more informed understanding which can add meaning to quantitative findings.

#### Limitations

Our study was limited in a single Midwestern geographical location therefore it is difficult to generalize results to a larger population. The study was also limited due to the lack of use of random sampling in which selection bias can occur with use of purposive sampling.

## Conclusion

Culturally adapted dietary interventions to improved eating practices among AA are needed to reduce the disproportionate impact ADRD has on the AA community. Older Midwestern’s perceived the MIND dietary model as most feasible and acceptable to AA culture. To enhance buy-in within the AA community, an adapted MIND dietary interventions need to incorporate salient AA cultural characteristics and strategies within both the design and delivery of the intervention.

## Supporting information

Table 1. Demographics of Study Participants

Table 2. Eating Habits

Table 3. Health Conditions Knowledge Interests

Table 4. Barriers to Eating Healthy

Table 5. Nutrition Education Preferences

Table 6. Themes, Subthemes, and Sample Quotes

## Data Availability

All data produced in the present work are contained in the manuscript.

## References

1. Association As. 2019 Alzheimer’s Disease Facts and Figures. Alzheimers Dement. 2019;15(3):321–97.

2. Morris MC, Tangney CC, Wang Y, et al. MIND diet slows cognitive decline with aging. Alzheimer’s & dementia : the journal of the Alzheimer’s Association. 2015;11(9):1015–1022. doi:10.1016/j.jalz.2015.04.011

3. Scarmeas N, Stern Y, Tang M-X, Mayeux R, Luchsinger JA. Mediterranean diet and risk for Alzheimer’s disease. Ann Neurol. 2006;59(6):912–921. doi:10.1002/ana.20854

4. Blumenthal JA, Smith PJ, Mabe S, et al. Longer Term Effects of Diet and Exercise on Neurocognition: 1-Year Follow-up of the ENLIGHTEN Trial. J Am Geriatr Soc. Mar 2020;68(3):559–568. doi:10.1111/jgs.16252

5. Epstein DE, Sherwood A, Smith PJ, et al. Determinants and consequences of adherence to the dietary approaches to stop hypertension diet in African-American and white adults with high blood pressure: results from the ENCORE trial. J Acad Nutr Diet. 2012;112(11):1763–1773. doi:10.1016/j.jand.2012.07.007

6. Airhihenbuwa CO, Kumanyika S, Agurs TD, Lowe A, Saunders D, Morssink CB. Cultural aspects of African American eating patterns. Ethnicity & Health. 1996;1(3):245–260.

7. Tangney CC, Kwasny MJ, Li H, Wilson RS, Evans DA, Morris MC. Adherence to a Mediterranean-type dietary pattern and cognitive decline in a community population–. The American journal of clinical nutrition. 2011;93(3):601–607.

8. Rankins J, Wortham J, Brown LL. Modifying Soul Food for the Dietary Approaches to Stop Hypertension Diet(dash) Plan: Implications for Metabolic Syndrome(dash of Soul). Ethnicity & disease. 2007;17(3):7–12.

9. Ard JD, Cox TL, Zunker C, Wingo BC, Jefferson WK, Brakhage C. A study of a culturally enhanced EatRight dietary intervention in a predominately African American workplace. J Public Health Manag Pract. Nov-Dec 2010;16(6):E1–E8. doi:10.1097/PHH.0b013e3181ce5538

10. Anderson-Loftin W. Soul Food Light: A Clinical Trial of a Culturally Competent Dietary Self-Management Intervention for Rural African-Americans With Diabetes. 2004:

11. Jefferson WK, Zunker C, Feucht JC, et al. Use of the Nominal Group Technique (NGT) to understand the perceptions of the healthiness of foods associated with African Americans. Evaluation and program planning. 2010;33(4):343–348.

12. Morris MC, Wang Y, Barnes LL, Bennett DA, Dawson-Hughes B, Booth SL. Nutrients and bioactives in green leafy vegetables and cognitive decline: Prospective study. Neurology. 2018;90(3):e214–e222. doi:10.1212/WNL.0000000000004815

13. Nilsson A, Salo I, Plaza M, Björck I. Effects of a mixed berry beverage on cognitive functions and cardiometabolic risk markers; A randomized cross-over study in healthy older adults. PLoS One. 2017;12(11):e0188173–e0188173. doi:10.1371/journal.pone.0188173

14. Williams JH, Auslander WF, de Groot M, Robinson AD, Houston C, Haire-Joshu D. Cultural relevancy of a diabetes prevention nutrition program for African American women. Health promotion practice. 2006;7(1):56–67.

15. Bisogni CA, Jastran M, Seligson M, Thompson A. How People Interpret Healthy Eating: Contributions of Qualitative Research. Journal of Nutrition Education and Behavior. 2012/07/01/ 2012;44(4):282–301. doi:https://doi.org/10.1016/j.jneb.2011.11.009

16. Story M, Kaphingst KM, Robinson-O’Brien R, Glanz K. Creating Healthy Food and Eating Environments: Policy and Environmental Approaches. Annual Review of Public Health. 2008/04/01 2008;29(1):253–272. doi:10.1146/annurev.publhealth.29.020907.090926

17. Brett J, Staniszewska S, Mockford C, et al. Mapping the impact of patient and public involvement on health and social care research: a systematic review. Health Expect. Oct 2014;17(5):637–50. doi:10.1111/j.1369-7625.2012.00795.x

18. Fortuna K, Barr P, Goldstein C, et al. Application of Community-Engaged Research to Inform the Development and Implementation of a Peer-Delivered Mobile Health Intervention for Adults With Serious Mental Illness. J Particip Med. Jan-Mar 2019;11(1):e12380. doi:10.2196/12380

19. Han H-R, Xu A, Mendez KJW, et al. Exploring community engaged research experiences and preferences: a multi-level qualitative investigation. Research Involvement and Engagement. 2021/03/30 2021;7(1):19. doi:10.1186/s40900-021-00261-6

20. Forsythe LP, Ellis LE, Edmundson L, et al. Patient and Stakeholder Engagement in the PCORI Pilot Projects: Description and Lessons Learned. J Gen Intern Med. Jan 2016;31(1):13–21. doi:10.1007/s11606-015-3450-z

21. Wennerstrom A, Springgate BF, Jones F, et al. Lessons on Patient and Stakeholder Engagement Strategies for Pipeline to Proposal Awards. Ethn Dis. 2018;28(Suppl 2):303–310. doi:10.18865/ed.28.S2.303

22. Dozier A, Hacker K, Silberberg M, Ziegahn L. Clinical and Translational Science Awards Consortium Community Engagement Key Function Committee Task Force on the Principles of Community Engagement. The value of social networking in community engagement http://www.atsdr….

23. Wallerstein NB, Duran B. Using community-based participatory research to address health disparities. Health promotion practice. 2006;7(3):312–323.

24. Krieger J, Allen C, Cheadle A, et al. Using community-based participatory research to address social determinants of health: lessons learned from Seattle Partners for Healthy Communities. Health Education & Behavior. 2002;29(3):361–382.

25. Champion VL, Skinner CS, Glanz K, Rimer BK, Viswanath K. Health behavior and health education. Theory, Research, and Practice (Eds). 2008:45–65.

26. Jones CL, Jensen JD, Scherr CL, Brown NR, Christy K, Weaver J. The Health Belief Model as an explanatory framework in communication research: exploring parallel, serial, and moderated mediation. Health Commun. 2015;30(6):566–576. doi:10.1080/10410236.2013.873363

27. Diddana TZ, Kelkay GN, Dola AN, Sadore AA. Effect of Nutrition Education Based on Health Belief Model on Nutritional Knowledge and Dietary Practice of Pregnant Women in Dessie Town, Northeast Ethiopia: A Cluster Randomized Control Trial. Journal of Nutrition and Metabolism. 2018/06/21 2018;2018:6731815. doi:10.1155/2018/6731815

28. McArthur LH, Riggs A, Uribe F, Spaulding TJ. Health Belief Model Offers Opportunities for Designing Weight Management Interventions for College Students. Journal of Nutrition Education and Behavior. 2018/05/01/ 2018;50(5):485–493. doi:https://doi.org/10.1016/j.jneb.2017.09.010

29. Birt L, Scott S, Cavers D, Campbell C, Walter F. Member Checking: A Tool to Enhance Trustworthiness or Merely a Nod to Validation? Qualitative Health Research. 2016/11/01 2016;26(13):1802–1811. doi:10.1177/1049732316654870

30. Tong A, Sainsbury P, Craig J. Consolidated criteria for reporting qualitative research (COREQ): a 32-item checklist for interviews and focus groups. Int J Qual Health Care. Dec 2007;19(6):349–57. doi:10.1093/intqhc/mzm042

31. Dedoose Version 9.0.17 web application for managing, analyzing, and presenting qualitative and mixed method research data. https://www.dedoose.com

32. Braun V, Clarke V. Using thematic analysis in psychology. Qualitative research in psychology. 2006;3(2):77–101.

33. Ismail HI, Chan KW, Mariod AA, Ismail M. Phenolic content and antioxidant activity of cantaloupe (cucumis melo) methanolic extracts. Food Chemistry. 2010/03/15/ 2010;119(2):643–647. doi:https://doi.org/10.1016/j.foodchem.2009.07.023

34. People H. Understanding and improving health. Washington, DC: US Dept of Health and Human Services; 2000.

35. Morland K, Wing S, Roux AD, Poole C. Neighborhood characteristics associated with the location of food stores and food service places. American journal of preventive medicine. 2002;22(1):23–29.

36. Gregory WH, Harper KW. The Ntu approach to health and healing. Journal of Black Psychology. 2001;27(3):304–320.

37. Woods-Giscombé CL, Black AR. Mind-body interventions to reduce risk for health disparities related to stress and strength among African American women: The potential of mindfulness-based stress reduction, loving-kindness, and the NTU therapeutic framework. Complementary health practice review. 2010;15(3):115–131.

