## Supplementary material for "Developing a culturally tailored brain healthy diet intervention for older midwestern African Americans: A Qualitative and Quantitative Study": Table 1. Demographics of Study Participants

| **Table 1. Demographics of study participants** | |
| --- | --- |
| **Gender** | |
| Male | 10 (25.6%) |
| Female | 29 (74.4%) |
| **Age** | |
| 55-64 | 11 (17.5%) |
| 65-75 | 16 (25.4%) |
| 75 years and older | 22 (34.9%) |
| **Marital Status** | |
| Single | 4 (10.3%) |
| Married | 21 (53.8%) |
| Widowed | 6 (15.4%) |
| Divorced | 8 (20.5%) |
| **Education** | |
| High School | 4 (10.3%) |
| Vocational/Trade | 1 (2.6%) |
| Some College | 7 (17.9%) |
| Associate | 3 (7.7%) |
| Bachelor | 9 (23.1%) |
| Masters | 13 (33.3%) |
| Doctoral | 2 (5.1%) |
| **Employment Status** | |
| Employed | 11 (28.2%) |
| Not employed | 1 (2.6%) |
| Retired | 27 (69.2%) |
| **Household Income** | |
| > $25,000 | 2 (5.1%) |
| $25,000 – $49,999 | 14 (35.9%) |
| $50,000 - $74,999 | 13 (33.3%) |
| $75,000 - $99,999 | 4 (10.3%) |
| $100,000 - $124,999 | 3 (7.7%) |
| $125,000 - $149,999 | 2 (5.1%) |
| $150,000 - $199,999 | 1 (2.6%) |
| **Number of adults in household not counting yourself** | |
| 0 | 13 (33.3%) |
| 1 | 18 (46.2%) |
| 2 | 7 (17.9%) |
| 3 | 0 |
| 4 | 1 (2.6%) |
| **Number of children under 18 in the household** | |
| 0 | 37 (94.9%) |
| 1 | 0 |
| 2 | 2 (5.1%) |
| Family history of dementia |  |
| Parent | 5 (12.8%) |
| Sibling | 2 (5.1%) |
| Other relative | 3 (7.7%) |
| **Health Conditions** | |
| Hypertension | 26 (66.7%) |
| High cholesterol | 20 (51.3%) |
| Diabetes | 8 (20.5%) |
| Overweight/obesity | 13 (33.3%) |
| **Religion** | |
| Christian | 37 (94.9%) |
| Other | 1 (2.6%) |
| Religiously Unaffiliated | 1 (2.6%) |
| **Religious Denomination** | |
| Baptist | 18 (48.6%) |
| Catholic | 1 (2.7%) |
| Methodist | 8 (21.6%) |
| Nondenominational | 8 (21.6%) |
| Pentecostal | 1 (2.7%) |
| Presbyterian | 1 (2.7%) |
