## Supplementary material for "Developing a culturally tailored brain healthy diet intervention for older midwestern African Americans: A Qualitative and Quantitative Study": Table 2. Eating Habits

| **Table 2. Eating Habits** | |
| --- | --- |
| **Current practices**  None  Low sodium  Low fat  Intermittent fasting  Other | (19, 48.7%)  (10, 25.6%)  (2, 5.1%)  Yes (8, 20.5%), No (31, 79.5%)  (6, 15.4%) |
| **Food consumption frequency**  Breakfast  Lunch  Dinner | 0-1 days/week (7, 17.9%), 2-3 days/week (8, 20.5%), 4-5 days/week (5, 12.8%), 6-7 days/week (19, 48.7%)  0-1 days/week (4, 10.3%), 2-3 days/week (5, 12.8%), 4-5 days/week (16, 41.0%), 6-7 days/week (14, 35.9%)  0-1 days/week (1, 2.6%), 2-3 days/week (1, 2.6%), 4-5 days/week (4, 10.3%), 6-7 days/week (33, 84.6%) |
| **Salt Intake**  Added Salt for cooking and preparation  Processed foods  Consumption of too much salt | Very Often (5, 12.8%), Often (6, 15.4%), Sometimes (12, 30.8%), Rarely (15, 38.5%), Never (1, 2.6%)  Very Often (3, 7.7%), Often (11, 28.2%), Sometimes (17, 43.6%), Rarely (7, 17.9%), Never (1, 2.6%)  Very Often (2, 5.1%), Often (3, 7.7%), Sometimes (15, 38.5%), Rarely (15, 38.5%), Never (4, 10.3%) |
| Food Preparation Practices | Baked (32, 82.1%), Air fried (17, 43.6%), Boiled (14, 35.9%), Fried (14, 35.9%), Grilled (18, 46.2%), Other (5, 12.8%) |
