## Supplementary material for "Developing a culturally tailored brain healthy diet intervention for older midwestern African Americans: A Qualitative and Quantitative Study": Table 3. Health Conditions Knowledge Interests

| **Table 3. Health Conditions Knowledge Interests** | |
| --- | --- |
| Alzheimer’s disease | Very Interested (19, 48.7%), Interested (16, 41.0%), Somewhat interested (3, 7.7%), Not interested at all (1, 2.6%) |
| Blood pressure | Very Interested (16, 41.0%), Interested (14, 35.9%), Somewhat interested (7, 17.9%), Not interested at all (2, 5.1%) |
| Cholesterol | Very Interested (14, 35.9%), Interested (13, 33.3%), Somewhat interested (10, 25.6%), Not interested at all (2, 5.1%) |
| Diabetes | Very Interested (14, 35.9%), Interested (15, 38.5%), Somewhat interested (7, 17.9%), Not interested at all (3, 7.7%) |
| Obesity | Very Interested (10, 25.6%), Interested (17, 43.6%), Somewhat interested (8, 20.5%), Not interested at all (4, 10.3%) |
