## Supplementary material for "Developing a culturally tailored brain healthy diet intervention for older midwestern African Americans: A Qualitative and Quantitative Study": Table 4. Barriers to Eating Healthy

| **Table 4. Barriers to Eating Healthy** | |
| --- | --- |
| Lack of time | 6 (15.4%) |
| Cost | 8 (20.5%) |
| Cooking Skills | 5 (12.8%) |
| Taste | 12 (30.8%) |
| Feeling Hungry | 6 (15.4%) |
| Other | 16 (41%) |
