## Supplementary material for "Developing a culturally tailored brain healthy diet intervention for older midwestern African Americans: A Qualitative and Quantitative Study": Table 5. Nutrition Education Preferences

| **Table 5. Nutrition Education Preferences** | |
| --- | --- |
| **Education Component Preferences**  Meal planning and prepping  Eating healthy on a budget  Food diaries  Reading food label  Nutrition content  Water intake | Very Interested (18, 46.2%), Interested (16, 41.0%), Somewhat interested (5, 12.8%), Not interested at all (0, 0.0%)  Very Interested (16, 41.0%), Interested (17, 43.6%), Somewhat interested (6, 15.4%), Not interested at all (0, 0.0%)  Very Interested (4, 10.3%), Interested (11, 28.2%), Somewhat interested (16, 41.0%), Not interested at all (8, 20.5%)  Very Interested (6, 15.4%), Interested (22, 56.4%), Somewhat interested (10, 25.6%), Not interested at all (1, 2.6%)  Very Interested (10, 25.6%), Interested (24, 61.5%), Somewhat interested (3, 7.7%), Not interested at all (2, 5.1%)  Very Interested (17, 43.6%), Interested (20, 51.3%), Somewhat interested (2, 5.1%), Not interested at all (0, 0.0%) |
| **Nutritionist or Health Education Coach Preferences**  In-person  By phone  Virtually | 35 (89.7%)  37 (94.9%)  37 (94.9%) |
| **Location Preference**  KU ADRC  Church  Virtually | 82.1%  37 (94.9%)  37 (94.9%) |
