## Supplementary material for "Developing a culturally tailored brain healthy diet intervention for older midwestern African Americans: A Qualitative and Quantitative Study": Table 6. Themes, Subthemes, and Sample Quotes

| **Table 6. Themes, Subthemes, and Sample Quotes** | | |
| --- | --- | --- |
| **Main Theme** | **Subtheme** | **Sample Quotes** |
| 1. Knowledge of dementia | 1.1 Family history    1.2 Impact on Black community/perceived cause of dementia    1.3 Role of diet in dementia | “I think that a healthy diet is important for every part of your body, especially the brain, because if we give the brain the correct nutrients, I just believe that we will, you know, function better and function longer if we eat the right things…you can’t fool the body.” |
| 2. Practices shaping food choices and consumption | 2.1 Household decision making  2.2 Food purchasing practices  2.3 Food preparation practices | “Since the pandemic, I have had….most of my shopping online and I go and pick it up, and I have a few smaller stores that I go to purchase things that um things like vegetables, things that I usually go in person to pick those out.” |
| 3. Barriers impacting healthy dietary consumption | 3.1 Access  3.2 Cost  3.3 Taste  3.4 Food spoilage | “Fruit and vegetables are expensive, and so whether you have, whether it's an Aldis, Aldis is probably cheaper than a lot of places, but there's not a lot of Aldis in some of the Black communities, it's not a lot that is accessible.” |
| 4. External instrumental support | 4.1 Cooking education  4.2 Accessibility guidance | “It also has to be available, it’s not like we can education, we can show, we can even provide taste, but we have to make it available and show them where they can obtain or acquire these healthy foods, these recipes, these meals. If we can show them easily how to get it, or how to maintain a healthy diet, try something once or twice, but I can’t include that into my everyday lifestyle rather easily if it’s going to be difficult, it’ll be more successful if it’s easier to obtain and maintain.” |
| 5. Elements of culturally tailored brain healthy dietary intervention | 5.1 Recruitment  5.2 Dietary model (MIND) (add seasoning/spice options, swap options, butter limitation barrier, like that its variety)  5.3 Delivery method  (cooking classes with samples, scriptures, recipes, collaboration w/ churches)  5.4 Retention (financial incentives, emphasis on short term & long-term benefits) | “I think having a sample there or a cooking class there to show people how you can use these products, and how much they cost, and have them actually taste it, you know they can see how to utilize it. They can see that it is good and healthy for them. I think that is a good key to sort of have people, so that the change, so they can change or maybe look at things differently.” |
